# Association of vitamin D level and maternal gut microbiome during pregnancy: Findings from a randomized controlled trial of antenatal vitamin D supplementation

**DOI:** 10.1101/2023.04.04.23288136

**Authors:** Andrea Aparicio, Diane R. Gold, Scott T. Weiss, Augusto A. Litonjua, Kathleen Lee-Sarwar, Yang-Yu Liu

## Abstract

Shifts in the maternal gut microbiome and vitamin D deficiency during pregnancy have been associated, separately, with health problems for both the mother and the child. Yet, they have rarely been studied simultaneously. Here, we analyzed gut microbiome (from stool samples obtained in late pregnancy) and vitamin D level (from blood samples obtained both in early and late pregnancy) data of pregnant women in the Vitamin D Antenatal Asthma Reduction Trial (VDAART), a randomized controlled trial of vitamin D supplementation during pregnancy, to investigate the association of vitamin D status on the pregnant women’s microbiome. To find associations we ran linear regressions on alpha diversity measures, PERMANOVA tests on beta diversity distances, and used the ANCOM-BS and Maaslin2 algorithms to find differentially abundant taxa. Analyses were deemed significant using a cut-off p-value of 0.05. We found that gut microbiome composition is associated with the vitamin D level in early pregnancy (baseline), the maternal gut microbiome does not show a shift in response to vitamin D supplementation during pregnancy, and that the genus *Desulfovibrio* is enriched in women without a substantial increase in vitamin D level between the first and the third trimesters of pregnancy. We conclude that increasing the vitamin D level during pregnancy could be protective against the growth of sulfate-reducing bacteria such as *Desulfovibrio*, which has been associated with chronic intestinal inflammatory disorders. More in-depth investigations are needed to confirm this hypothesis.

## Introduction

Vitamin D has been strongly associated with many conditions of human health. Initially it was discovered to promote bone health [1,2] but now it is known to have many other physiological functions among which, importantly, are modulating immunological and inflammation responses [3–5]. Vitamin D can be obtained from supplements, diet or sun exposure and is converted by the liver into its serum form 25-hydroxyvitamin D which, in turn, is converted into its activated form 1,25-dihydroxyvitamin D [6] by the kidneys and many other cells in the body. The biological activity of vitamin D’s active form in the body is mediated by the vitamin D receptor (VDR), primarily expressed in the intestines, kidneys, parathyroid gland and bones, though also expressed by several cells of the immune system [7]. In the gut, a growing amount of evidence indicate that vitamin D and VDR signaling are responsible for maintaining epithelial integrity and modulating immune and inflammatory responses when the barrier is penetrated by bacteria, leading to chronic states of inflammation when the signaling is disrupted [6,8].

The gut microbiome is composed of trillions of microbes that collectively influence the health of its host [9]. In particular, the gut microbiome is known to have important immunoregulation functions [10] and disruptions of it are associated with chronic inflammatory diseases [11,12]. Strong evidence that vitamin D is associated with microbiota alterations exists, but the results are considerably heterogeneous [13–15].

The gut microbiome composition changes with age and is naturally modified during several stages of life. During pregnancy, the microbiome shift often resembles diseased states or dysbiosis that, surprisingly, instead of leading to a decline in fitness, promotes homeostasis [16]. The gut microbiome is dramatically modified between the first and third trimesters of pregnancy and bacteria related to inflammation and immune response processes are usually elevated as the pregnancy progresses [17,18].

The impact on the offspring of maternal vitamin D level in serum and of vitamin D consumption during pregnancy has been studied several times, establishing associations with the children’s microbiome and with outcomes such as changes in the immune system modulation, asthma/allergic disease incidence, likelihood of *C. difficile* colonization, and changes in microbiome richness and composition [19–21]. However, to our knowledge, no clinical trials have focused on the impact of vitamin D consumption on the mothers’ own microbiome during pregnancy. One observational study found an association between vitamin D dietary intake and a reduction of microbiome richness, and an increase of bacteria with pro-inflammatory properties [22], however, due to the lack of serum vitamin D level measurements, and considering that its absorption may depend on many factors other than only diet, the knowledge gap persists. Changes in the maternal gut microbiome are known to be associated with the development of preeclampsia, gestational diabetes and other serious pregnancy pathologies [23] and with negative offspring outcomes, especially with those related to immunity or allergic diseases [24–26]. Thus, exploring the effects of maternal vitamin D status on their microbiome is not only useful to identify potential associated pregnancy complications or impacts on the mothers’ health. Additionally, it has been repeatedly shown that the maternal and infant microbiome are related [24,25,27,28] and, therefore, this direction of research can also contribute to discover potential consequences for the children’s health.

Here we analyze the 16S rRNA data generated from stool samples of 114 pregnant women in the Vitamin D Antenatal Asthma Reduction Trial (VDAART), a multi-site randomized, double-blind, placebo-controlled trial of vitamin D supplementation during pregnancy, to look for associations between the participants’ vitamin D status and their gut microbiome during the third trimester of pregnancy. We use three different measures for vitamin D status: (1) baseline vitamin D level; (2) treatment assignment; and (3) change in vitamin D level over the trial period. In the following, we present three key findings. First, the baseline vitamin D level is associated with the gut microbiome composition. Second, the gut microbiome is robust enough such that the vitamin D supplementation (even with up to 4,400 IU cholecalciferol daily) during pregnancy does not significantly modify it. Third, *Desulfovibrio,* a genus of Gram-negative sulfate-reducing bacteria, is enriched in women whose vitamin D level did not have a substantial increase (less than10ng/mL) over the trial period.

## Methods

### Vitamin D Antenatal Asthma Reduction Trial (VDAART)

VDAART is a randomized controlled trial of Vitamin D supplementation during pregnancy to prevent asthma in offspring conducted in the United States (St. Louis, Boston, and San Diego; NCT00920621). 870 women were enrolled at 10-18 weeks of gestation and randomized to receive either a high dose of vitamin D oral supplements (4,400 IU cholecalciferol daily, called *treatment or high dose treatment* hereafter) or a *placebo* dose of vitamin D oral supplements (400 IU cholecalciferol daily) between enrollment and delivery. The study protocol was approved by the institutional review boards at each participating institution and at Brigham and Women’s Hospital. All participants provided written informed consent [29].

### Stool sample collection and processing

During the third trimester of pregnancy (weeks 32-38 gestation), 120 participants of the VDAART trial provided a stool sample. Subjects were asked to collect a 0.5 teaspoon-sized sample 1 to 2 days before a study visit and store the sample in a home freezer before transport with a freezer pack to the study site. Stool was not collected if participants had used antibiotics in the past 7 days. After delivery to the study site, stool samples were immediately stored at - 80°C. Microbiome profiling was performed by sequencing the 16S rRNA hypervariable region 4 (V4 515F/816R region) on the Illumina MiSeq platform at Partners Personalized Medicine (Boston, MA). Of the 120 stool samples collected, 2 with fewer than 1,000 reads measured were excluded from the analysis, leaving 118 samples.

### Blood sample collection and processing

Maternal blood samples were collected at two time points: at enrollment at 10-18 weeks of gestation and at 32-38 weeks of gestation. The serum 25-hydroxyvitamin D 25(OH)D level (termed as *vitamin D level* hereafter) was measured in both samples using the DiaSorin LIAISON method (a chemiluminescence assay) at the Channing Division of Network Medicine, Brigham and Women’s Hospital (Boston, MA). All the 118 women who provided a stool sample in the third trimester of pregnancy had provided a blood sample at the first time point (yielding a baseline vitamin D level) and 114 of them also provided one at the second time point (yielding a *final* vitamin D level). Therefore, we have blood samples to analyze the change in vitamin D level between early and late pregnancy of 114 women; 59 of them were on the treatment group and the remaining 55 were on the placebo group.

### Serum vitamin D level

For the 114 women with baseline and final measurements (Figure 1.a–b), we calculated the vitamin D level *difference* as the final level value minus the baseline level value; the mean vitamin D level difference among the 114 women was of 11.2 ng/mL (Figure 1.c). We used a rounded cut-off of 10 ng/mL to categorize the women into two groups: *high change* (difference of more than 10 ng/mL in serum 25hydroxyvitamin D 25(OH)D) and *low change* (difference equal to or less than 10 ng/mL in serum 25-hydroxyvitamin D); 62 women fell into the high change group and 52 in the low change group. Additionally, we created a variable that simultaneously accounts for the change of vitamin D level and the baseline vitamin D level (see SI for details and corresponding analysis).

**Figure 1:**
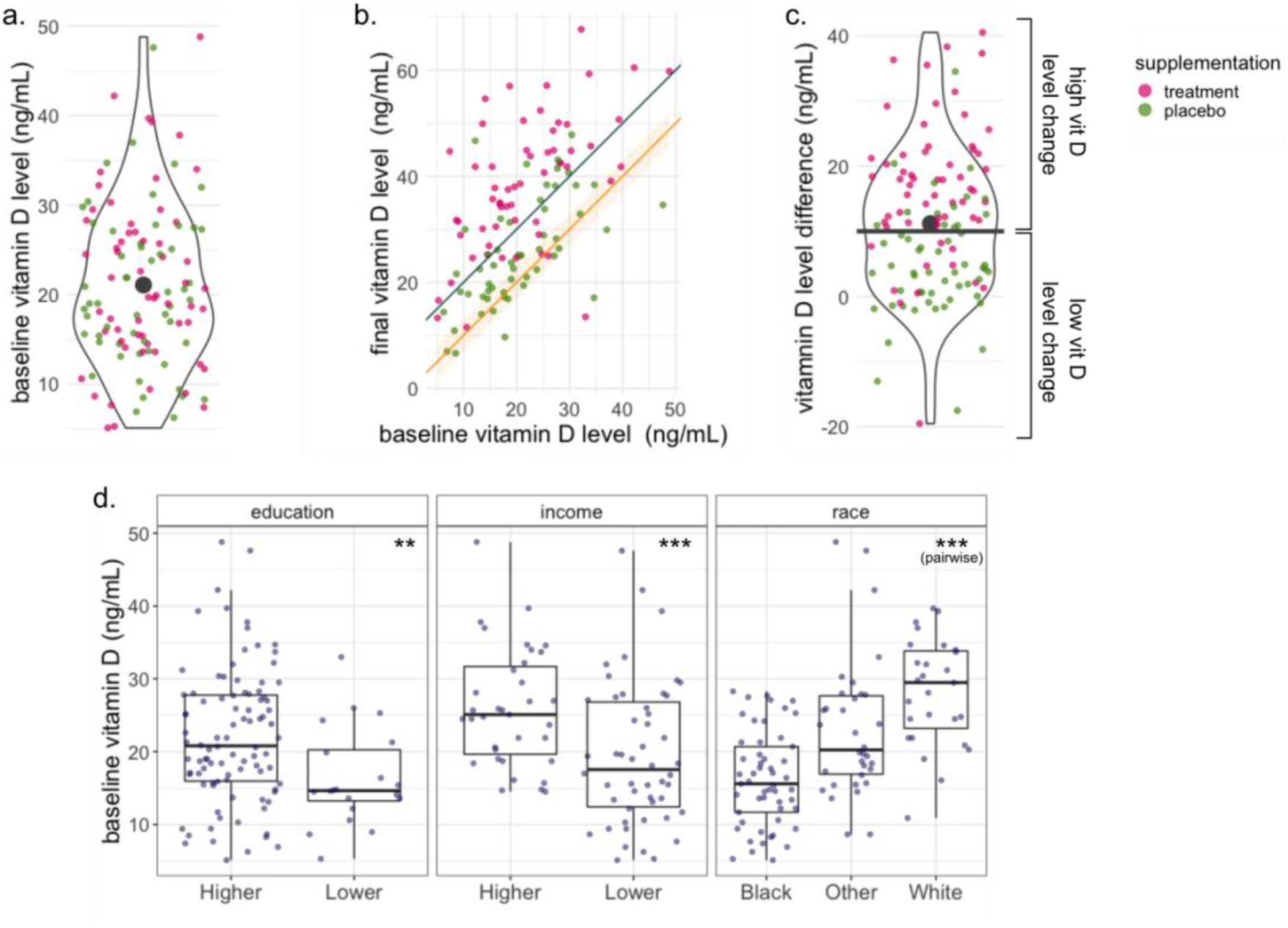
Vitamin D status was measured in three different ways: baseline vitamin D level (serum 25-hydroxyvitamin D 25(OH)D [ng/mL] measured in a blood sample at enrollment), treatment assignment (treatment or placebo vitamin D supplementation), and vitamin D level change over the trial period (high change or an increase of at least 10 ng/mL between the enrollment and third trimester, and low change or a difference of less than 10 ng/mL). **a.-c.** The colors of the markers in the three panels represent the treatment assignment of every participant. **a**. Distribution of baseline vitamin D level of all the participants. The black marker represents the mean baseline vitamin D level in all participants (21.01 ng/mL). The treatment assignment is not associated to the baseline vitamin D level. **b.** Baseline vitamin D level vs final vitamin D level. The orange line and band trace a difference of 0±3 ng/mL between the baseline and the final measurements. The gray line traces a difference of 10 ng/mL between baseline and final measurements. Most of the participants had an increase in their vitamin D level over the trial period. **c**. Distribution of the Vitamin D level difference over the trial period. The black marker represents the mean vitamin D difference in all participants (11.27 ng/mL); the black horizontal line represents the cut-off of 10ng/mL that separates the high and low change groups. **d.** Distribution of the participants’ baseline vitamin D level stratified by the different characteristics. The baseline vitamin D value is significantly associated with the participants’ education, income, and race.

### Mothers’ characteristics

The stool and blood samples were collected at three different sites: Boston, MA; San Diego, CA, and St. Louis, MI. The available subjects’ characteristics from the initial enrollment questionnaire are the mothers’ age, race, education, household income, and history of asthma and hay fever. Mothers were categorized into groups for each factor except for their age which was kept as a continuous variable. Race and ethnicity information was collected because they are determinants of circulating 25-hydroxyvitamin D levels; the race and ethnicity of every participant was self-reported. Participants were asked to first categorize themselves as either Hispanic or non-Hispanic, then to categorize their race into prespecified categories. Race/ethnicity (called *race* hereafter) groups were collapsed into 3 groups for the analysis: Black or African American mothers (called *Black mothers* hereafter), White, non-Hispanic mothers (called *White mothers* hereafter), and Hispanic mothers and mothers of other race (n=25 and n=9, respectively, called *Other race* hereafter). Education groups: higher education (some college education or more, n=94) and lower education (technical school, high-school, or less, n=20). Income groups: higher income (household income of more than 50 000 U.S. dollars/year, n=35) and lower income (less than 50 000 U.S. dollars/year, n=50). History of asthma or hay fever: yes (n=39, n=65, resp.) or no (n=65, n=49, resp).

### Statistical Analyses

For the statistical analyses we measured vitamin D status in three different ways: baseline vitamin D level (measured at enrollment), randomized treatment assignment (treatment or placebo supplementation), and vitamin D level *change* between baseline and final measurements (high or low change). Unless specifically noted, we used a cut-off p-value of 0.05 to deem results statistically significant for all our analyses. All analyses of microbiome associations were adjusted for two *a priori* selected potential covariates: participants’ race and education level, unless otherwise specified.

### Alpha Diversity

Alpha diversity measures are estimates of an individual sample’s taxonomic diversity. We computed the observed richness, Shannon, and Simpson indices using the *Phyloseq* package in R [30]. The observed richness simply counts the number of different taxa present in each sample. The Shannon and Simpson indices incorporate the measures of richness and evenness of every sample. We used Wilcoxon rank-sum tests and covariate adjusted linear regressions to compare the samples’ estimates of these three alpha diversity metrics by measures of vitamin D status.

### Beta Diversity

Beta diversity measures quantify the dissimilarity in taxonomic composition between two samples. We computed the following measures: Bray-Curtis dissimilarity, Jaccard distance, Unifrac distance, and Weighted-Unifrac distance using the *Phyloseq* package in R. We used the adonis2 algorithm in the R package *vegan* [31], which performs PERMANOVA tests using the distance matrices, to find associations between the beta-diversity dissimilarity measurements and measures of maternal vitamin D status.

### Abundance Association Analysis

To ensure robustness of the identified associations, we used two different methods, and their corresponding R packages, that look for associations between the subjects variables and the abundance of specific taxa: *ANCOM-BC* [32] and *MaAsLin* [33]. ANCOM-BC is a differential abundance method that uses a log-linear regression framework with a sample-specific offset term to account for differences in sampling fraction between samples. MaAsLin is a multivariate association method that uses additive linear models to detect associations between specific groups and the abundance of taxa simultaneously treating all the present taxa as outcomes. We used the *false discovery rate* (FDR) method to adjust the p-values for multiple comparisons. The association analyses were performed on genus-level data.

## Results

### Subject characteristics

The mean baseline vitamin D level in early pregnancy among all 114 participants was 21.08 ng/mL (Figure 1.a). Table 1 summarizes the participants’ characteristics with respect to their baseline vitamin D level. We found a significant association between the baseline vitamin D level and the mothers’ race, income, and education level (ANOVA p-value *=* 3.6 × 10^−9^, 2.8 × 10^−4^, 0.01, respectively). Black women tended to have the lowest baseline vitamin D level, White women tended to have the highest, and Latino/Hispanics and other women of other races tended to be in between; women with lower income and women with lower education tended to have lower levels of baseline vitamin D (Figure 1.d). The race and education variables were included as covariates in all downstream analyses, while the household income was excluded from the covariates due to high missingness (29 women, i.e., 25% of our cohort did not provide income data) and because it is strongly correlated with maternal education and race (Chi-squared test p-value < 7 × 10^−3^, 5 × 10^−4^, respectively). Additionally, we found a significant association between the collection site and the women’s baseline vitamin D level (ANOVA p-value < 0.001). The collection site is correlated with the subjects’ race (Chi-squared test p-value < 0.0005) and thus it was also excluded from the covariates. The baseline vitamin D level was treated as a continuous variable in all the downstream analyses except for the differential abundance algorithm ANCOM-BC, for which we used the mean baseline vitamin D level of 21.01 ng/mL as a cut-off to define two groups: *below the mean* and *above the mean*.

**Table 1:**
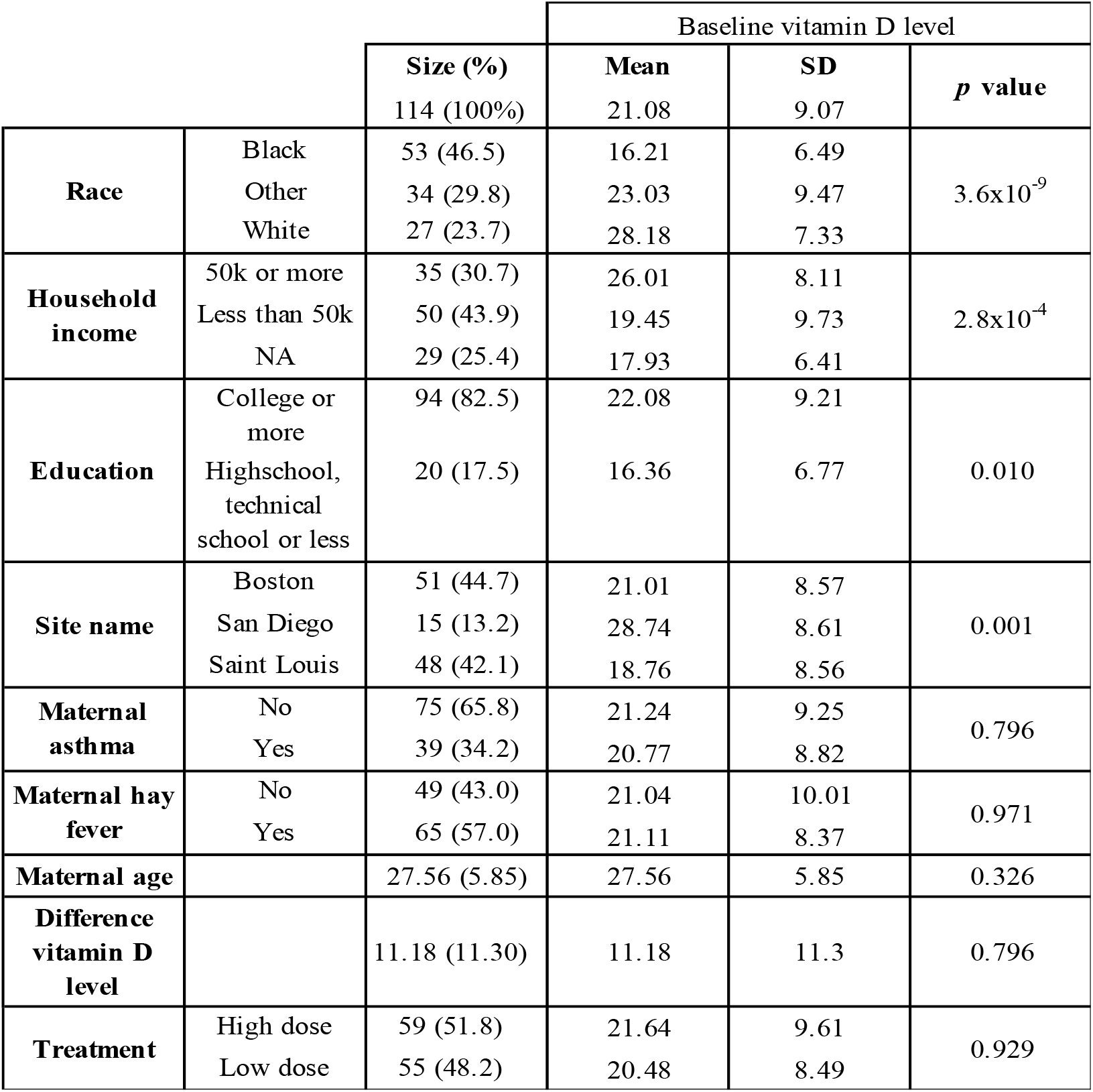
Summary of the participants’ characteristics with respect to their measurement of Serum 25-hydroxyvitamin D 25(OH)D level at enrollment (baseline vitamin D level).

The participants of VDAART were randomized to receive either a high (4400 IU daily cholecalciferol) or a low (400 IU daily cholecalciferol) dose of vitamin D supplements (called *treatment* or *placebo*, respectively, hereafter) [34,35]. Table 2 summarizes the participants’ characteristics stratified by their treatment assignment, revealing that the randomization performed for the original VDAART cohort (>800 women) resulted in similar baseline characteristics between treatment groups among the 114 women with baseline and final vitamin D measurements.

**Table 2:** Summary of the participants’ characteristics with respect to the treatment group they were assigned to.

|  |  |  | Treatment assignment |  |  |
| --- | --- | --- | --- | --- | --- |
|  |  | Overall | Treatment | Placebo | *p* value |
|  |  | 114 | 59 | 55 |  |
| Race | Black | 53 (46.5) | 26 (44.1) | 27 (49.1) | 0.614 |
|  | Other | 34 (29.8) | 20 (33.9) | 14 (25.5) |  |
|  | White | 27 (23.7) | 13 (22.0) | 14 (25.5) |  |
| Household income (%) | 50k or more | 35 (30.7) | 19 ( 32.2) | 16 ( 29.1) | 0.927 |
|  | Less than 50k | 50 (43.9) | 25 ( 42.4) | 25 ( 45.5) |  |
|  | NA | 29 (25.4) | 15 ( 25.4) | 14 ( 25.5) |  |
| Education (%) | College or more | 94 (82.5) | 48 ( 81.4) | 46 ( 83.6) | 0.941 |
|  | Highschool, technical school or less | 20 (17.5) | 11 ( 18.6) | 9 ( 16.4) |  |
| Baseline vitamin D level (mean (SD)) |  | 21.08 (9.07) | 21.64 (9.61) | 20.48 (8.49) | 0.496 |
| Site name (%) | Boston | 51 (44.7) | 29 ( 49.2) | 22 ( 40.0) | 0.543 |
|  | San Diego | 15 (13.2) | 8 ( 13.6) | 7 ( 12.7) |  |
|  | Saint Louis | 48 (42.1) | 22 ( 37.3) | 26 ( 47.3) |  |
| Maternal asthma (%) | No | 75 (65.8) | 39 ( 66.1) | 36 ( 65.5) | 1.000 |
|  | Yes | 39 (34.2) | 20 ( 33.9) | 19 ( 34.5) |  |
| Maternal hay fever (%) | No | 49 (43.0) | 26 ( 44.1) | 23 ( 41.8) | 0.958 |
|  | Yes | 65 (57.0) | 33 ( 55.9) | 32 ( 58.2) |  |
| Maternal age (mean (SD)) |  | 27.56 (5.85) | 27.47 (5.55) | 27.65 (6.19) | 0.871 |

Most of the 114 women had an increased vitamin D level between early and late pregnancy, though the extent of vitamin D increase varied between participants (Figure 1.a). The mean difference between the baseline and final vitamin D level among the participants was 11.2 ng/mL. A cut-off of 10 ng/mL in vitamin D level difference yielded two groups: high vitamin D level change (increase of at least 10 ng/mL) with 62 subjects, and low vitamin D level change (decrease in vitamin D or increase of less than 10ng/mL) with 52 subjects. We did not find any significant associations between the change groups and the participants’ characteristics, suggesting that the vitamin D level change was likely not confounded by them (Table S2). The vitamin D level in women in the treatment group (n=59) increased by 16.8 ng/mL on average (48 of them fell in the high change group and 11 of them in the low change group), while the vitamin D level in the women in the placebo group (n=55) increased by only 5.11 ng/mL on average (14 of them fell into the high change group and 41 of them in the low change group). The treatment assignment is significantly associated with the vitamin D level change assignment (Chi squared test p-value < 6.7 × 10^−9^, Figure S3.b). A significant vitamin D level difference between the treatment groups, and the absence of an association with the baseline vitamin D level were also reported for the entire VDAART cohort [34]. We found that the vitamin D level change was not associated with the baseline vitamin D level (Figure S3.a). To further reduce the risk of confounding, we ran all the downstream analysis for a variable baseline-change combination variable, with no qualitative changes to the results (SI).

### Microbiome composition is associated with baseline vitamin D level

We found a small but not statistically significant positive correlation between the baseline vitamin D level and gut microbiome richness or for the three alpha diversity indices analyzed (Figure 2.a). We found a significant association between the women’s baseline vitamin D level and their microbiome composition for all beta diversity measurements (Bray-Curtis, Jaccard, Unifrac and W-Unifrac adonis2 (PERMANOVA) p-values *<* 0.025, 0.025, 0.015, and 0.045, respectively). This association was not modified in models adjusted for education and race (Bray-Curtis, Jaccard, Unifrac and W-Unifrac adjusted adonis2 (PERMANOVA) p-values *<* 0.015, 0.015, 0.015, and 0.035, respectively). A covariate adjusted (education and race) linear regression between every pair of subjects’ difference in baseline vitamin D and the corresponding beta diversity distance between them confirmed the significant association (p-value *<* 2 × 10^−16^ for all distances, Figure 2.b). However, specific individual taxa associated with baseline vitamin D were not identified by either the ANCOM-BC or MaAsLin2 algorithms adjusted for both education and race (all FDR > 0.05 at the genus level).

**Figure 2:**
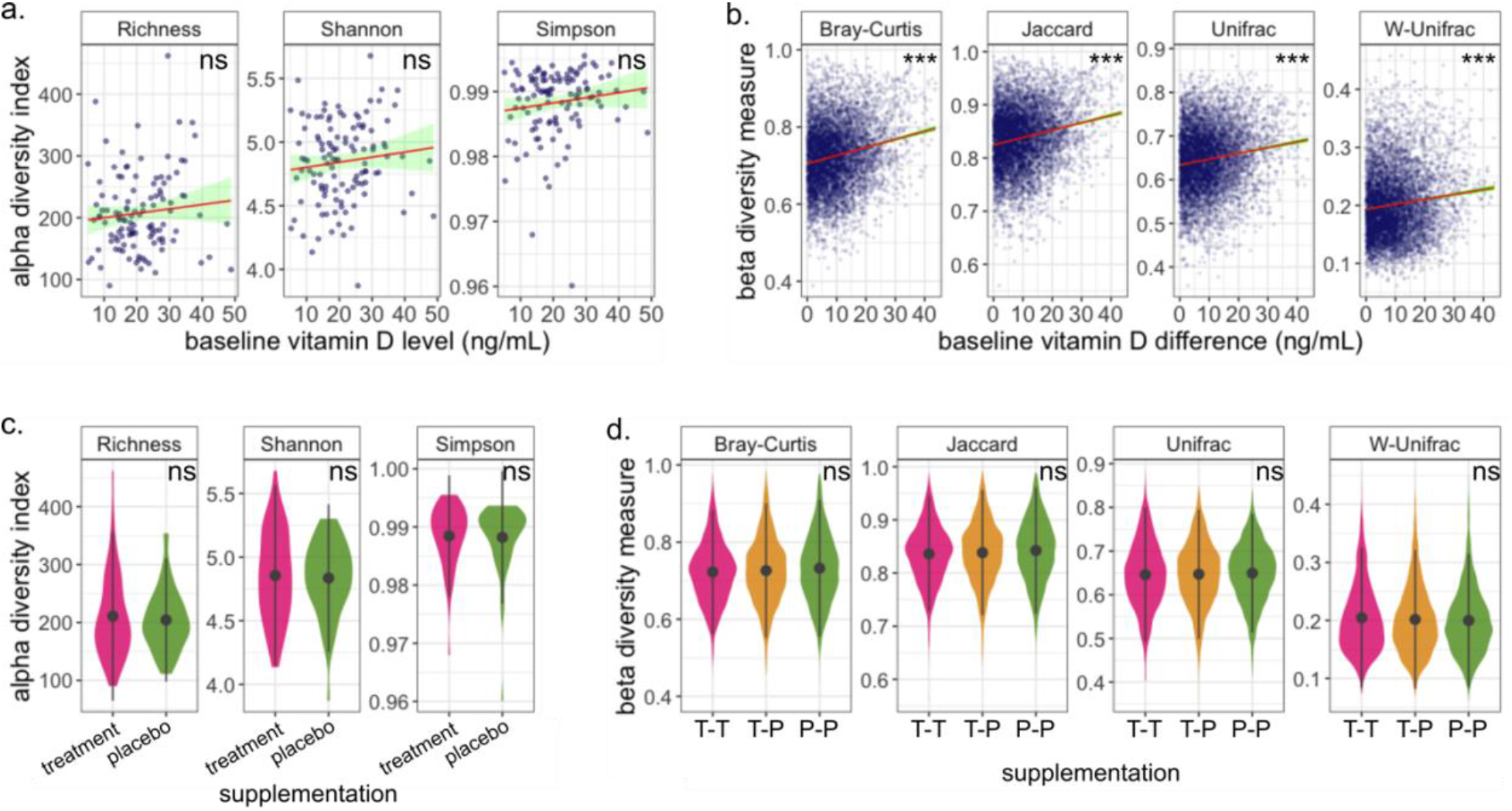
The baseline vitamin D is associated with the microbiome composition but not with its richness; the microbiome is robust to vitamin D supplementation. **a.-b.** The red line traces a linear model fit of the panel’s data and the green band represents a 95% confidence interval. **a.** The markers represent every individual’s baseline vitamin D level with respect to their alpha diversity. No significant association between the baseline vitamin D level and the alpha diversity indices was found. **b.** Every marker represents the difference between the baseline vitamin D levels of two participants with respect to their corresponding beta diversity measurement. We found a significant association between the baseline vitamin D level and all the beta diversity indices that we analyzed (Bray-Curtis, Jaccard, Unifrac and Weighted-unifrac). Pairs of participants with a larger difference in baseline vitamin D level tend to have more dissimilar microbiome composition than pairs whose baseline vitamin D level is closer to each other. **c.-d.** the black markers represent the mean values, and the black vertical lines represent the standard deviation. **c.** Distribution of the alpha diversity indices among the two treatment groups. We found no association between the microbiome richness and the subjects’ treatment assignment for any of the analyzed alpha diversity indices analyzed (Observed, Shannon and Simpson diversity). **d.** The magenta (green) violins represent the distribution of inter-individual gut microbiome dissimilarity of subjects assigned to the high dose of vitamin D supplementation group (placebo group), labeled as T-T (P-P). The yellow violins represent the distribution of gut microbiome dissimilarity between subjects assigned to the treatment group and those assigned to the placebo group, labeled as T-P. There is no association between the treatment assignment and any of the dissimilarity measures.

### Gut microbiome is robust to vitamin D supplementation during pregnancy

In an intention-to-treat analysis, we found no significant associations between the participants’ treatment assignment and any of the alpha diversity indices that we calculated (Figure 2.c), suggesting that the vitamin D supplementation did not modify the participants’ microbiome diversity during the trial period. We also did not find any significant associations with the analyzed beta diversity measurements. (Figure 2.d), suggesting that the microbiome composition was not altered with the short-term treatment during pregnancy. Additionally, both differential abundance algorithms, ANCOM-BC and MaAsLin2, failed to identify any taxa that was significantly differentially expressed between the groups (all FDR > 0.05). This suggests that the microbiome during pregnancy is robust to short-term vitamin D supplementation over the treatment period.

### Change in vitamin D level does not impact microbiome diversity

Comparing participants by the change in their vitamin D level allows for assessment of the impact of vitamin D over a relatively short period (between week 10-18 and week 32-38 of pregnancy) on the microbiome, and accounts for potential lack of adherence to the vitamin D supplementation treatment assignment. All the vitamin D level change analyses were adjusted for the participants’ baseline vitamin D level in addition to their race and education. We found no significant associations between the participants’ vitamin D change and their microbiome richness for any of the alpha diversity indices that we analyzed. We also found no significant associations with the women’s microbiome composition for any of the analyzed beta diversity measurements. This suggests that the microbiome richness and composition are robust to changes in the vitamin D level during pregnancy.

### *Desulfovibrio* is enriched in pregnant women with low change of vitamin D level

Next, we looked at differentially abundant taxa between the change groups at the genus level. We found that two genera were enriched in the low change group (*Clostridium sensu stricto 1* and *Desulfovibrio*, ANCOM-BC p-value < 0.04 and 0, respectively), and that two genera were depleted (*Coprococcus* and *Fusicatenibacter*, ANCOM-BC p-value < 0.02 for both) (Figure 3.a). However, after adjusting for multiple comparisons, only *Desulfovibrio* remained significantly differentially abundant (ANCOM-BC FDR < 0.01, log fold change = 0.0064). MaAsLin analysis also revealed an enrichment in *Desulfovibrio* in women with low change in vitamin D (coefficient of 0.04 and p-value *<* 8 × 10^−5^, q-val < 0.01, blue line in Figure 3.b). Indeed, *Desulfovibrio* was expressed almost exclusively in participants that had a low change of vitamin D level (area on the left of solid gray line in Figure 3.b), and it was not expressed in any of the participants whose vitamin D level increased by 15 ng/mL or more (area on the right of the discontinuous line in Figure 3.b). Finally, we found significant associations between the relative abundance of *Desulfovibrio* and (Figure 3.c) the participants’ history of asthma (linear model adjusted for education level coefficient of 0.0045 and p-value < 0.016), education, and income, (Wilcoxon rank sum test p-val *<* 0.03, for both). We found a weak association between the abundance of *Desulfovibrio* and the participants’ race (ANOVA p-value=0.671) and no significant association with treatment assignment. In particular, *Desulfovibrio* was not present in stool samples from any White participant and was less abundant in participants with higher income and education compared to those with lower income and education. However, the significant associations between the abundance of *Desulfovibrio* and vitamin D change were preserved after adjusting the ANCOM-BC and MaAsLin algorithms for race and education as covariates. Noteworthy, a linear model adjusted for education level showed that there is no significant association between Desulfovibrio and the mothers’ baseline vitamin D level, but analyses simultaneously accounting for the change of vitamin D level and the baseline vitamin D level pointed toward an enrichment of *Desulfovibrio* among participants with a low increase in vitamin D over pregnancy (see SI for detailed results).

**Figure 3:**
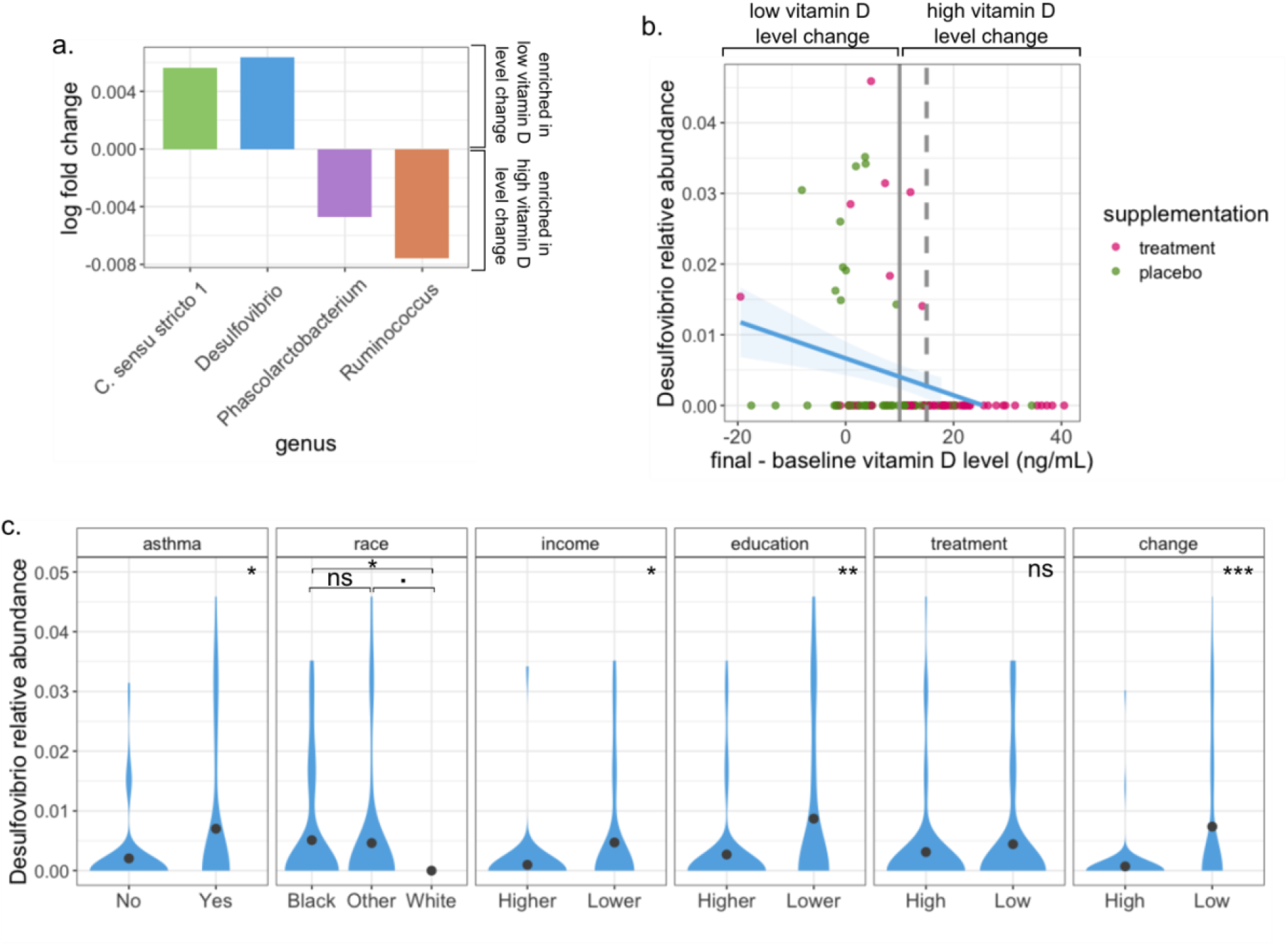
The sulfate-reducing bacteria *Desulfovibrio* is overexpressed in subjects with low vitamin D level change. **a**. Log fold change of differentially abundant genera. Two genera are enriched in the low change group, and two are depleted (significant ANCOM-BC p-value). When adjusting for multiple comparisons, the association remains significant only for the overexpression of *Desulfovibrio* (significant ANCOM-BC q-value). **b** Vitamin D level change (final-baseline measurement) vs *Desulfovibrio* relative abundance. The vertical solid line represents the 10 ng/mL cut-off that defines the high and low change groups. *Desulfovibrio* is almost exclusively present in participants with low vitamin D level change (left of solid vertical line) and is absent in those that had a vitamin D level difference larger than 15 ng/mL (discontinuous vertical line). The blue line traces a lineal model regression fit with 95% confidence interval (blue transparency). **c.** *Desulfovibrio* relative abundance distribution with respect to participants’ characteristics. A significant association exists between *Desulfovibrio*’s abundance and the subjects’ history of asthma, race, income, education, and vitamin D level change. No significant association with the subject’s treatment assignment was found. *Desulfovibrio* was not present in White women.

Interestingly, we found that the presence of *Desulfovibrio* is negatively, yet weakly, correlated with a low consumption of fruits, when adjusted for education level (maternal diet summary derived from the principal component analysis of an 18-item food frequency questionnaire data. The second principal component represents a diet low in citrus and other fruits. Adjusted linear regression p-value = 0.0826). Additionally, we found that *Desulfovibrio* was present in a very small proportion (1.6%, n=4) of the offspring gut microbiomes in 244 children who provided a stool sample between 3 and 6 months of age. None of the infants with a presence of *Desulfovibrio* were born to White mothers, mothers with higher income or had developed asthma at 3 or 6 years old.

## Discussion

Here we described the vitamin D status in pregnant women in three different ways (baseline vitamin D level, randomization to high or low dose vitamin D supplementation, and vitamin D level change over the trial period), each one shedding light on the relationship with the gut microbiome in its own way.

The baseline vitamin D level (measured at enrollment at 10-18 weeks of gestation) provides a perspective of the mothers’ background in terms of their vitamin D status. For example, it might reflect their eating habits, history of supplementation, exposure to the sun, and/or ability to process the acquired vitamin D through any of these mechanisms. The identified association between the participants’ baseline vitamin D level and their race and education level is in agreement with previous findings that indicate a higher risk of vitamin D deficiency in non-Whites [36,37] and in individuals with a lower education level [36]. Also, lower income, which is strongly associated with lower education level and non-White ethnicity in our data, is known to be associated with lower vitamin D intake. We found a small and not statistically significant positive correlation between the baseline vitamin D level and the alpha diversity, and a significant association between the baseline vitamin D level and the subjects’ microbiome composition. The latter has been associated with the development of allergies, asthma or other respiratory diseases [38]. Because an adequate vitamin D level has been iteratively found to be protective against airway inflammation processes [39], immune-mediated disorders [4,40], and inflammation [5], our finding may help fill in some of the mechanistic gaps in the microbiome-vitamin D-asthma axis.

VDAART was originally designed to evaluate the impact of prenatal maternal vitamin D supplementation on asthma and respiratory diseases in children [29,34,35] but it also provides an opportunity to evaluate other effects of the treatment assignment. We found that while most mothers’ vitamin D level increased over the trial period, especially so for those on the treatment group, their microbiome composition seems to not be affected by the treatment assignment. This suggests that vitamin D supplementation during pregnancy might be beneficial for the mothers in protecting them against vitamin D deficiency-related disorders and, potentially, for their offspring [21,22,41] without impacting the mothers’ risk with respect to microbiome shifts during the pregnancy period. However, depending on the chronicity of vitamin D deficiency prior to pregnancy, there may be physiologic adaptations to deficiency that may need longer periods of supplementation prior to pregnancy to effect change in the mothers’ vitamin D levels. Consequently, and based on our finding of an association between baseline vitamin D level and microbiome composition, in order for changes in the microbiome to be consistently detected across different microbiome-related downstream analysis, a longer supplementation period in addition to longer periods between measurements might be needed.

Vitamin D level in people under a supplementation schedule not only depends on their assigned dose, but can be determined by a multitude of factors such as their vitamin D level at baseline, sunlight exposure, diet, the liver’s efficiency in converting the absorbed vitamin D into its serum form, adherence to the treatment, the amount of time their vitamin D level has been in a (in) sufficient range, certain health conditions, or genetic predisposition [42]. Therefore, the change in vitamin D level over a certain period allows a more accurate vision of its associations with the microbiome than information on vitamin D intake via supplements or diet. We found enrichment of the genus *Desulfovibrio* in subjects whose vitamin D level had a low change over the trial period*. Desulfovibrio* is the most common sulfate-reducing bacteria in the human intestine [43], and is also found in water sediments and environments with extreme temperature or pH [44]. In humans, the presence of *Desulfovibrio* has been associated with an increased incidence of inflammatory bowel diseases [45,46], and may even have a causal role in their pathophysiology [47,48]. Our result might indicate that an adequate vitamin D level can be protective against a higher risk of developing IBD or other related conditions by way of modulating abundance of this gut microbe. This result agrees with previous studies that found negative correlations between *Desulfovibrio’s* abundance and vitamin D dietary [49] and supplements [50] intake. Moreover, the inability to absorb and process vitamin D has been found to cause dysbiosis and the development of sulfate-induced colitis in mice [51]. However, this pathway requires further clarification; for example, our data contradicts the result of an observational study that found a positive correlation between vitamin D dietary intake and *Desulfovibrio* abundance [52]. Interestingly, some evidence of negative correlations between *Desulfovibrio* and respiratory ailments is available [53,54], but we found a positive correlation with the mothers’ history of asthma. On the other hand, we found that no children in the VDAART trial with asthma had a presence of *Desulfovibrio* in their gut microbiome, but the genus was found in only a tiny proportion of the subjects. Thus, more studies in this direction are necessary to establish a conclusive association. Our data revealed an increased prevalence of *Desulfovibrio* in participants of more disadvantaged backgrounds (lower income and education level), as well as an important difference between the race groups. To our knowledge, this link has not been explored in depth before, and studies that aim to address this gap would help establish an association.

Some limitations of this study are the relatively small sample size, short trial duration, lack of metagenomic sequencing and the lack of stool microbiome data from the 1^st^ trimester of pregnancy. The small sample size could potentially decrease the generalizability of our results, although we do have a diverse cohort from geographically distant study sites. The duration of the study, both in terms of the supplementation duration and the time between baseline measurements and follow up, might hinder from detecting changes in the microbiome more robustly. 16s rRNA sequencing offers many advantages over shotgun metagenomic sequencing such as the lower cost, sensitivity to contamination and bioinformatic requirements, with the drawback that is limited to the 16s region of the rRNA gene only, leading to a loss in resolution in terms of taxonomic detection capabilities. While 16s rRNA sequencing has been the most prevalent genetic marker for decades, full gene sequencing data would allow a more in-depth analysis of the role that vitamin D plays in the microbiome during pregnancy. Finally, the availability of gut microbiome data at enrollment would provide a more accurate comparison of the vitamin D supplementation effects on the microbiome by establishing a baseline microbiome status for every participant.

In summary, here we have shown evidence that suggests that the vitamin D level in serum is linked to the gut microbiome composition in pregnant women, but not to its richness. Also, that the gut microbiome seems to be robust to vitamin D supplementation during pregnancy, but that an increase in vitamin D level between the first and third trimesters of pregnancy, in addition to all its other well-known health benefits, could protect against the growth of sulfate-reducing bacteria, such as *Desulfovibrio*, which is associated to several respiratory and bowel inflammation-related ailments.

### Authors’ contributions

STW, KLS, and YYL designed research; AA, KLS, and YYL conducted research; DRG, and AAL, provided essential databases, AA performed statistical analysis; AA wrote paper with input from KLS and YYL; AA, KLS, and YYL had primary responsibility for final content; all authors have read and approved the final manuscript.

### Data sharing

Microbiome sequencing data from VDAART are part of the ECHO consortium and ECHO consortium members can obtain the data directly from the ECHO DCC or for those not part of ECHO directly from the authors. All other relevant data are available from the authors upon reasonable requests.

### Code sharing

Code used to generate the results is available from the authors upon requests.

## Data Availability

All data produced in the present study are available upon reasonable request to the authors

